# Analysis of a deeply-phenotyped familial hypercholesterolemia cohort from Mexico shows a role for both rare and common alleles across known dyslipidemia genes and reveals structural variation in a novel locus

**DOI:** 10.1101/2025.04.24.25325891

**Authors:** Nicholas Katsanis, Niki Mourtzi, Consuelo D. Quinto-Cortés, Alexandro J. Martagon, Alexander G. Ioannidis, Francisco M. De La Vega, Jeff Gulcher, Ming Ta Michael Lee, Mohammad A. Faghihi, Arturo Lopez-Pineda, Sonia Moreno-Grau, Daniel Mas Montserrat, Míriam Barrabés, David Bonet, Pavel Salazar Fernandez, Jeff Wall, Babak Moatamed, Roopa Mehta, Gabriela A. Galan-Ramirez, Rafael Zubirán, Daniel Elias-Lopez, FH Mexican Registry Group, Teresa Tusié-Luna, Carlos A. Aguilar-Salinas, Carlos D. Bustamante

**Author notes:** Authors contributed equally (Nicholas Katsanis, Niki Mourtzi, Consuelo D. Quinto-Cortés). Corresponding authors. **Address correspondence to: Dr. Carlos D. Bustamante** & **Dr. Nicholas Katsanis** Galatea Bio, Inc., 14350 Commerce Way, Miami Lakes, Florida 33016, United States of America; **Dr. Carlos A. Aguilar-Salinas** Instituto Nacional de Ciencias Médicas y Nutrición Salvador Zubirán, Vasco de Quiroga 15, Belisario Domínguez Secc 16, Tlalpan, 14080, Mexico City, Mexico.

## Abstract

Familial hypercholesterolemia (FH) is a genetic disorder driven in part by mutations in three genes that encode components of the cholesterol pathway: *LDLR*, *APOB*, and *PCSK9*. However, the majority of FH genetics has been performed in individuals of European descent. Here, we leveraged a cohort of 300 patients from the Mexican FH registry to understand how rare, high liability alleles and common variants might contribute to shaping individual risk. Using a combination of whole exome and of short- and long-read whole genome sequencing, we report three key findings. First, we observed that rare pathogenic point mutations and structural variants in all known FH genes, together with variants in *APOE*, *CREB3L3*, and *PLIN1*, contribute to a molecular FH diagnosis in 67% of families, including novel gene-disruptive CNVs which arose in a native American background. Second, ancestry-adjusted polygenic risk score analysis identified a significant liability for coronary artery disease, hypertension, LDL, HDL, and Type 2 Diabetes. The polygenic signal for LDL was present in patients with rare, pathogenic FH mutations and was more prominent in individuals bereft of a molecular FH diagnosis. Finally, we report both a whole-gene duplication and common, non-coding variants in a novel locus, *PDZK1*, which contribute to the genetic burden of FH, a finding we replicated in the UK Biobank (UKB). Together, our analyses illustrate the value of genetic studies in non-European populations and reinforce the notion that individual risk to disease can arise from both rare, large effect alleles (alone or in combination across genes) and common variants that increase the mutational burden of a biological system.

## Introduction

Familial hypercholesterolemia (FH) is a disorder characterized by high levels of low-density lipoprotein cholesterol (LDL-C) that contribute significantly to early-onset cardiovascular disease **[1]**. FH has a strong genetic component, with heterozygous mutations in *LDLR*, *APOB* and *PCSK9* found in 60-80% of clinically-diagnosed individuals [1] and transmitted as a dominant trait with reduced penetrance and variable expressivity **[2][3]**. In rare cases, recessive mutations have been associated with a severe form of the disease, known as ‘homozygous FH’ (HoFH). This form of the disease presents with extreme LDL levels during infancy or childhood, xanthomas (cholesterol deposits in the skin or tendons), premature atherosclerosis, and a high frequency of cardiovascular disease [4].

FH remains underdiagnosed and undertreated, with some estimates placing the disease burden at 14-34 million people worldwide [5]. Despite the magnitude of impact and the subsequent burden of cardiovascular disease to individuals, families, and national health systems, strategies geared toward early diagnosis and management of the disorder are lacking, with genetic studies often applied retrospectively, after adult-onset clinical disease has impacted cardiovascular health [6].

A meta-analysis of 19 epidemiological studies conducted between 1990 and 2017 that used either clinical or genetic criteria to diagnose FH in an aggregate cohort of ∼2.5M people estimated a population incidence of 1:250 [7], doubling previous estimates of 1:500 **[8]**, and highlighting the need to implement prophylactic screening measures for an actionable disease. Other estimates of the prevalence of FH have been even higher, with incidence rates of 1:200 reported **[9]**. Some of the variance in disease incidence is likely caused by differences across populations. Most early FH estimates were derived from individuals of European descent [5][10] Subsequent studies from Japan [11], Iran [12], Brazil[13], and Norway[14] and others have not only provided evidence for population-specific differences in incidence rates, but have also shown that some deleterious variants are likely enriched or unique in some ancestral backgrounds. Although such findings improve our understanding of the genetic architecture of FH around the world, they also highlight the challenge of interpreting the pathogenic potential of hitherto unknown alleles. In that context, the Clinical Genome Resource Familial Hypercholesterolemia (FH) Variant Curation Expert Panel [15] has refined the standardized variant classification guidelines created by the American College of Medical Genetics and Genomics (ACMG) and the Association for Molecular Pathology (AMP) **[16]**. Such efforts are clearly necessary, yet their success in driving accurate interpretation and, ultimately, diagnosis, hinges on the availability of deeply phenotyped, genetically diverse, FH patients.

The Mexican FH registry (www.fhmexico.org.mx) was launched in 2017 with a mandate to help close the FH knowledge gap in the country’s population which has significant admixture of European, Native American and African individuals [17]. As part of this work, the Registry has collected >300 families with FH and has generated detailed profiles of clinical manifestation and progression, while also performing targeted genetic studies [18]. Here, we present the first comprehensive genetic analysis of this cohort: coupling whole exome and short- and long-read whole genome sequencing with state-of-the art ancestry and ancestry-adjusted polygenic score algorithms, we have studied the contribution of both point mutations and structural variants in all genes associated with dyslipidemias in humans. Moreover, we have examined how the genetic architecture of FH might be influenced by the concomitant contribution of rare and common alleles. Finally, we have asked whether some of the missing genetic burden of FH might be contributed either by novel alleles (in known FH genes) unique to this population or in novel disease-associated loci.

## Materials and Methods

### Recruitment and clinical environment

The FH Mexican Registry design and rationale has been described [17]. A web-based registry was created to collect data on individuals with heterozygous/homozygous familial hypercholesterolemia (FH), with clinicians from private and public institutions invited to participate. To date, 60 institutions from 28 of 32 Mexican states are involved in the FH Mexican Registry [17]. This research protocol has been approved by the Institutional Research and Ethics Committees (Ref. 2571). Patients identified with baseline LDL-C levels >190 mg/dL (adults) or >160 mg/dL (children) are evaluated in accordance with the Dutch Lipid Clinic Network criteria [5]. Only those cases that have a definite or probable diagnosis are included. Decision to participate in the study and informed consent form (ICF) are obtained from each subject. Within the web-based registry information is captured from subjects with heterozygous/homozygous familial hypercholesterolemia (FH) clinical diagnosis. This data is in accordance with the European Atherosclerosis Society (EAS) FH recommendations. In the registry, each patient is assigned a unique code in order to preserve anonymity. Follow-up data on each patient is also collected once a year, or in accordance with their normal clinical follow-up.

### Sequencing

Whole exome sequencing (WES) libraries were prepared with the Twist library Preparation EF Kit 2.0 with Twist Exome 2.0/Twist SNP Diversity Panel in a 4/1 ratio (Twist Biosciences, San Francisco, CA, USA) according to manufacturer’s protocol. Briefly, genomic DNA was fragmented enzymatically, ligated to a universal adapter and amplified with Twist Unique Dual Index Primers. Amplified samples were then hybridized overnight with the Twist 2.0 and Diversity panels. The hybridized fragments were sequenced as 2 x 150 bp reads on an Illumina NovaSeq 6000 instrument (Illumina, San Diego, CA, USA). The exome (283,942 regions) and the SNP diversity panel (1,380,542 SNPs) were sequenced at an approximate mean depth of 50X and 10X coverage, respectively. In addition, a subset of samples carrying putative copy number variants or pathogenic missense variants were sequenced by both Illumina short-read WGS and Nanopore sequencing. Nanopore sequencing was done as follows: Sequencing libraries were prepared from 1 ug of DNA using the Oxford Nanopore Technologies (ONT) Ligation Sequencing kit V14 (SQK-LSK114). The samples were loaded on the PromethION Flow Cells R10 (M Version) and were wash and reloaded after approximately 24 hours. The samples were sequenced for a total of 72 hours on a PromethION 24 sequencer. Mapping, alignment, and variant calling of the long reads was performed with Sentieon DNAscope software (Sentieon, Inc., San Jose, CA, USA).

### Variant calling and Quality Control

We generated VCF files with the Illumina DRAGEN™ Secondary Analysis (Version 4.2.4), which we filtered using default parameters, including a quality score < 10.41 for SNVs, a quality score < 7.83 for indels, a read depth ≤ 1, and the removal of genotype calls inconsistent with the expected chromosome ploidy. To confirm survey-based family architecture, we used Plink v1.9 [19] to perform a sex check and to calculate relatedness in all pairs of samples in our cohort. -. Finally, we imputed variants using the GLIMPSE2 algorithm with a reference panel from 4,091 individuals from the 1000 Genomes Project and the Human Genome Diversity Project, as implemented by Gencove Inc. (New York, NY, USA).

### Ancestry and PRS

We used, G-Nomix [20] to calculate local and global ancestry proportions. Our ancestry model was trained to detect genomic loci with the following ancestries: Native American, European, African, South East Asian, Central and South Asian, East Asian, Levantine and Middle Eastern, and Oceanian. We also used the Galatea Bio PRS, which includes an ancestry-adjusted PRS algorithm, to estimate the genetic risk of high LDL-C levels for all the samples in the cohort. The algorithm performs ancestry adjustment by normalizing the score with a mean and standard deviation computed using a neighborhood of genetically-close individuals from a proprietary database of diverse samples covering populations across the world. As controls, we used an - internal database of individuals from Mexico with no evidence of FH diagnosis. To reduce potential biases originated by using a population-level control, for each sample in our FH cohort, we select the closest individual (in the PCA space) of our population-level control cohort in order to generate a subset of control individuals with an ancestry composition and genetic structure similar as possible as the FH cohort. PRS for LDL-C levels were calculated using the variants and weights of the Galatea Bio LDL PRS model.

### Mutation analysis

#### a. SNV annotation

Initially, we focused our analysis on genes known to cause FH: *LDLR*, *APOB*, *PCSK9* and *LDRLAP1*. We annotated variants using Variant Effect Predictor (VEP) [21] and filtered for population-based gnomAD allele frequency less than 1% (https://gnomad.broadinstitute.org/). In addition to the classical genes, FH cases with pathogenic variants in *APOE* have been reported, suggesting that this locus should also be included in genetic screening for FH [22]. Next, we examined non-classical genes implicated either with FH or with conditions mimicking FH; we searched for rare and likely pathogenic (per modified ACMG criteria) variants in these genes (**Table S1**).

We also performed AI-based prioritization and scoring of candidate disease genes and diagnostic conditions with the Fabric Enterprise GEM algorithm (Fabric Genomics, Inc., Oakland, CA USA)[23], which uses - variant files and patient’s HPO terms as input and combines variant prioritization algorithms and clinical/genomic database annotations using Bayesian scoring. - The algorithm adjusts its parameters based on proband-specific variant data and uses naive Bayes to integrate factors such as ancestry, gene location, inheritance patterns, and proband sex to refine variant scoring. Furthermore, variants of interest were classified according to the ACMG/AMP guidelines [16] with the aid of Fabric ACE automated variant classification engine, adding manual curation for literature-based criteria to obtain a final classification.

#### b. CNV calling and validation

To call Copy Number Variation (CNVs) we used the WES read depth data. We ran CNV calling using two algorithms designed for WES data: Codex [24] and ExomeDepth [25] with the recommended default settings of each program. Briefly, to generate the normalized coverage metrics for each exon, we supplied as inputs the BED files of our target gene set and the BAM files processed in batches according to each WES run. CODEX normalizes the depth of coverage by applying a Poisson latent factor model that removes bias due to factors such as GC content, exon capture and amplification efficiency, and latent systemic artifacts. Five Poisson latent factors were included in the normalization model for this dataset. For each sample, ExomeDepth built a reference set that included samples with similar read count distribution and then performed CNV calling. We prioritized CNVs detected by both programs.

We confirmed candidate CNVs (deletions and duplications) involving multiple exons with custom-made TaqMan assays (ThermoFisher Scientific,Waltham, MA, USA). Single-exon CNVs were validated orthogonally against CNV calls derived from WGS data from the suspected cases. For WGS data, we employed the Illumina DRAGEN Secondary Analysis v4.2 CNV/SV caller with default parameters, which integrates sequencing depth and split-read analyses [26]. The DRAGEN pipeline has shown superior sensitivity and specificity compared to other CNV callers used in WGS data [27].

### Breakpoint analysis

We generated WGS data for three samples with candidate CNVs: a carrier of a whole gene duplication in *PCSK9* and two carriers of an exon13-14 *LDLR* deletion. By using the sequencing reads of the WGS we identified the breakpoints of each CNV event using the Manta algorithm [28]. Next, we extracted the region surrounding the breakpoints (+-1000 bp on each side) and searched for repetitive elements using RepeatMasker [29]. The repetitive elements at the left and right breakpoints were then compared for similarity with BLASTn [30].

### Analysis for genes participating in cholesterol pathway

In addition to all known dyslipidemia genes, we also studied rare variants (SNVs and CNVs) in all known genes that encode components of the cholesterol pathway. In particular, we searched for likely deleterious variants in 47 genes defined to participate in cholesterol biosynthesis and transport (KEGG pathway: hsa048979) along with their interactors (55 genes) extracted from the Intact database [31] **(Table S1.**)

### CNV calling in UK Biobank

To replicate novel variants of interest with respect to their involvement in hypercholesterolemia, we examined the cholesterol profiles of UK Biobank (UKB) participants in the UK [32]. Copy number variant (CNV) detection was performed using PennCNV version 1.0.5 [33], with log R ratio and B-allele frequency (BAF) data provided by the UK Biobank, setting missing values to NA. Across the cohort, CNVs on chromosome 1 were analyzed separately for each of the 106 genotyping batches, each containing approximately 4,700 samples. Batch-specific population frequency of the B allele (PFB) files were generated by calculating the mean BAF for each batch. Probes with missing PFB values were excluded in a batch-specific manner. The hidden Markov model (HMM) file for the Affymetrix genome-wide 6.0 array, included in the PennCNV-Affy package, was used directly for analysis [34].

## Results

### Cohort demographics, phenotypes, and genetic structure

We studied 170 families from the Mexican FH National Registry [17]. Of these, 111 were composed of a single affected individual with an available DNA sample, while in 59 families we were able to phenotype and recruit multiple members across several generations; we focused on individuals with a “definite” or “probable” FH diagnosis as defined by the Dutch Lipid Clinic Network Score (DLCNS), which includes LDL-C levels >190mg/dl in adults and 160mg/dl in children [17]. A summary of cohort characteristics is shown in **Fig. 1 and Table 1**, where we recorded standard anthropometric data; lipid profiles (including secondary dyslipidemia phenotypes such as xanthomas or corneal arcus); and other relevant phenotypic data such as the incidence of stroke, coronary heart disease, and Type 2 Diabetes.

**Figure 1:**
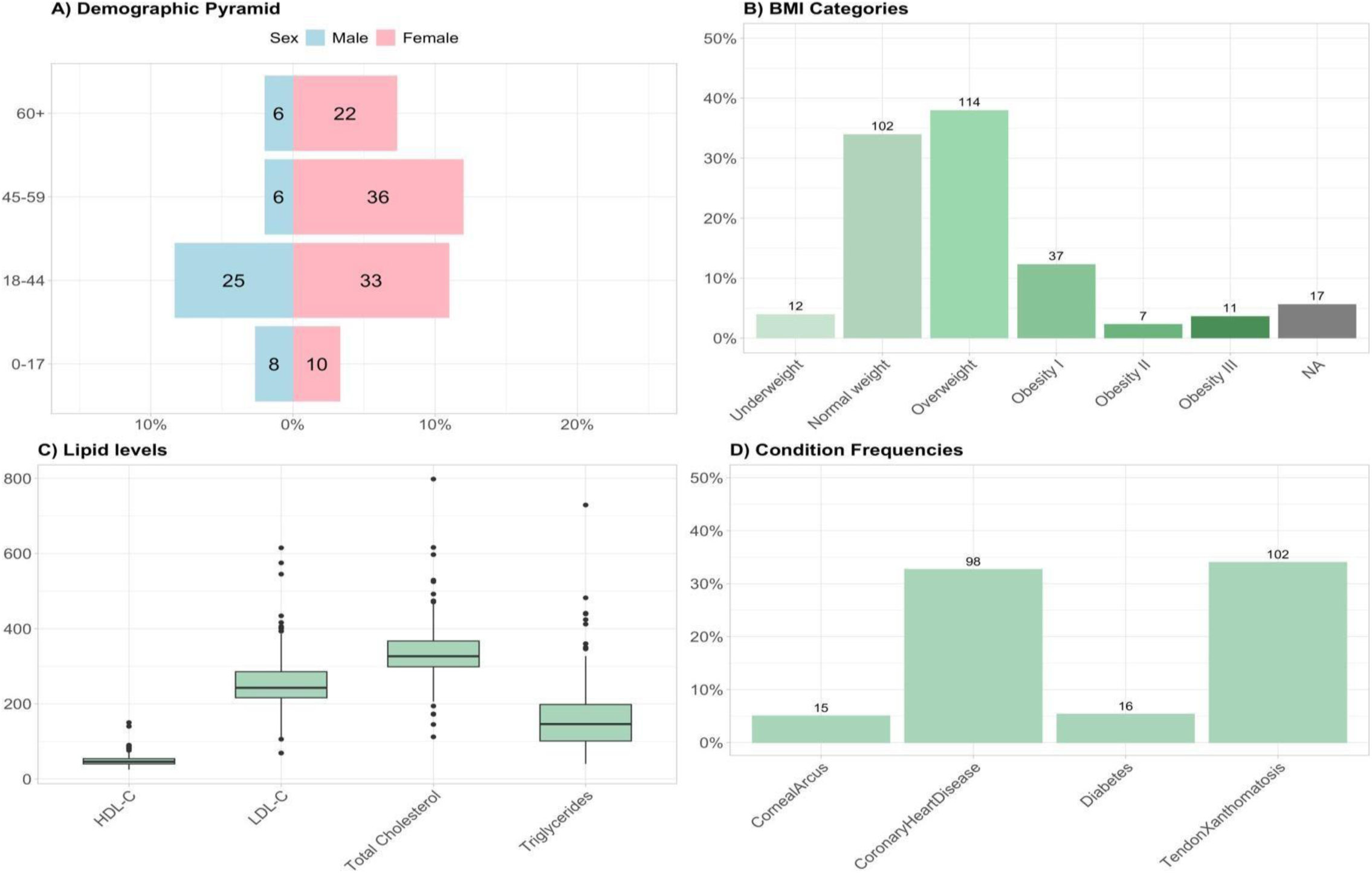
Overview of Cohort Characteristics. A) demographic pyramid that displays the age and sex distribution of participants; B) Distribution of BMI categories across the cohort; C) Distribution of lipids: total cholesterol, LDL, HDL, and triglycerides D) Distribution of health conditions: corneal arcus, coronary heart disease, diabetes and tendon xanthomatosis.

**Table 1:** Demographic and clinical characteristics of the FH cohort.

|  | **Overall** |
| --- | --- |
| **N** | 300 |
| **Sex** |  |
| Female | 201 (67%) |
| Male | 99 (33%) |
| **Age** |  |
| Age at FH diagnosis,<br>mean (s.d.) | 42.24 (±18.89) |
| **BMI** |  |
| weight/height<sup>2</sup>, mean<br>(s.d.) | 26.35 (±5.40) |
| **Current or past smoker** |  |
| Current, n (%) | 29 (13.8%) |
| Past smoker, n (%) | 10 (4.8%) |
| Never smoker, n (%) | 171 ( 81.4%) |
| **Corneal arcus** |  |
| Yes, n (%) | 15 (7.2%) |
| No, n (%) | 194 (92.8%) |
| **Xanthoma** |  |
| Yes, n (%) | 102 (34%) |
| No, n (%) | 194 (92.8%) |
| **Diabetes** |  |
| T2D, n (%) | 16 (7.6%) |
| None, n (%) | 194 (92.4%) |
| **Coronary heart disease** |  |
| Yes, n (%) | 98 (46.7%) |
| No, n (%) | 112 (53.3%) |
| **Stroke** |  |
| Yes, n (%) | 5 (2.4%) |
| No, n (%) | 205 (97.6%) |
| **Total cholesterol** |  |
| mmol/L, mean (s.d.) | 336.28 (±80.32) |
| **LDL-cholesterol** |  |
| mmol/L, mean (s.d.) | 254.96 (±76.61) |
| **HDL-cholesterol** |  |
| mmol/L, mean (s.d.) | 49.11 (±16.42) |
| **Triglycerides** |  |
| mmol/L, mean (s.d.) | 162.92 (±95.26) |

Seventy-seven individuals received targeted genetic testing for *LDLR*, *APOB* and *PCSK9*, the data from which have been reported [35] but we pursued a cohort-wide, comprehensive analysis of all LDL-relevant genes and signatures by performing joint Illumina short-read WES and low-pass genotype-by-sequencing (GBS) of ∼1.4M Diversity Panel SNPs. QC showed that >98% of the WES-targeted regions were covered at a minimum of 20x, whilst the diversity SNP set achieved 2-10x coverage for ∼80% of sites. We excluded five samples from further analyses: three were proven to be duplicates and two were of low quality, leaving us with data for 300 individuals from 167 families.

We first assessed all survey-based familial relationships. Identity-by-descent analysis confirmed kinships for 278/300 individuals. Eight individuals did not map to any family and were thus reassigned as singletons, while the remaining 14 individuals were found to be 1^st^ or 2^nd^ degree relatives of existing multi-member families and were reassigned as such.

Next, we determined eight-label ancestries for each individual in our study (**Fig. 2 / Table 2**.). Consistent with prior genetic analyses from this geographic region [36], 6 individuals showed nearly 100% Amerindian (n=5) or European (n=1) ancestry, while the -remained of the cohort were admixed between these two ancestral groups. We did not observe any significant representation from other ancestries, with the exception of ∼10 individuals whose genome contained 10% of more segments of East Asian, African or Middle Eastern origin. This pattern has been observed in similar admixed samples from Mexico, derived from the three-way admixture process that took place after the arrival of Europeans to the continent [37].

**Figure 2:**
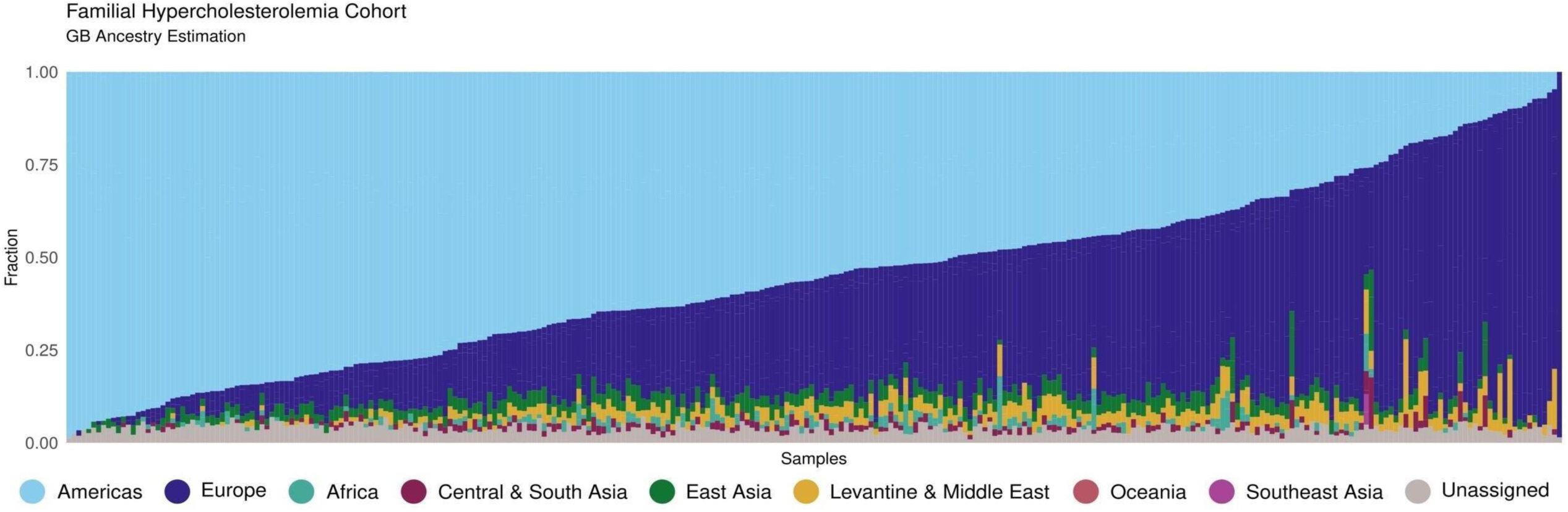
Ancestry analysis. Each bar corresponds to an individual and the height of each color within the bar indicates the proportion of the individual’s genome that can be attributed to the corresponding ancestral population. The “unassigned” fraction of the genome represents regions that could not be confidently assigned to any specific ancestry.

**Table 2:**
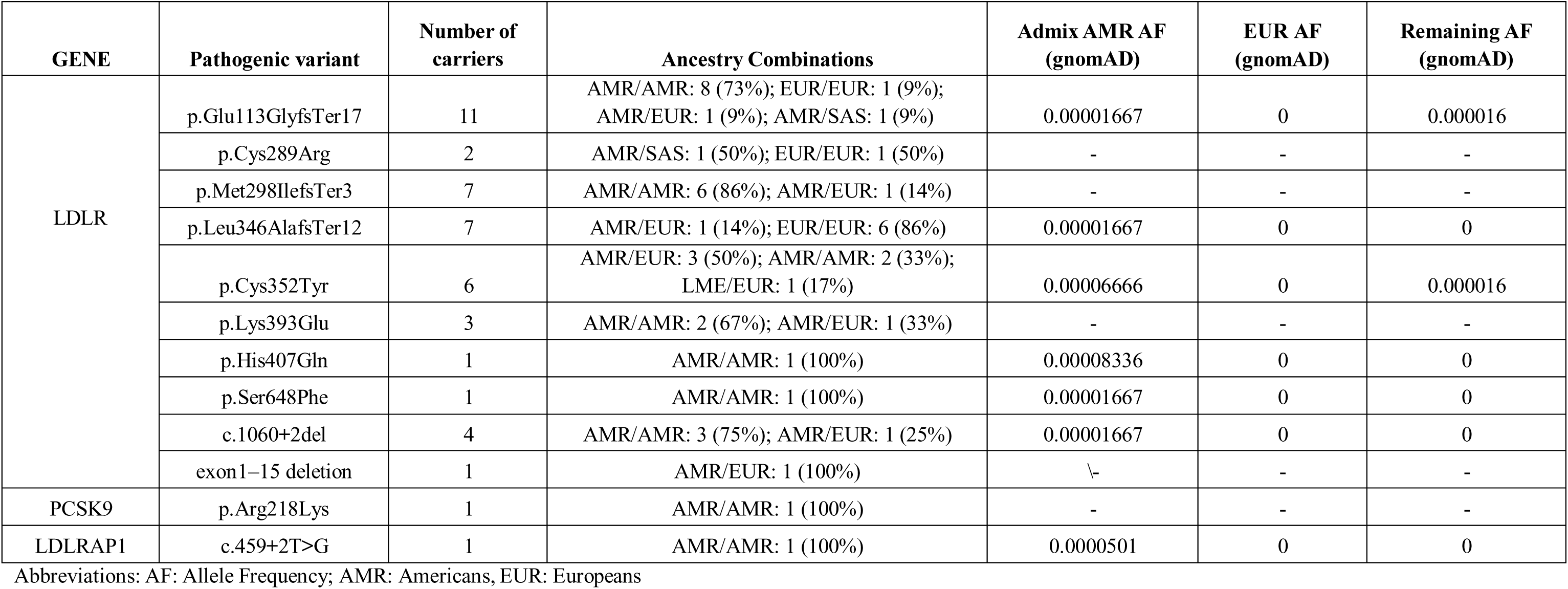
Ancestry analysis for samples carrying ultra-rare or unreported variants in gnomAD.

### Analysis of known FH genes

As a first step toward mapping the genetic architecture of FH in this cohort, we interrogated all rare variants in the three “classical” genes dominant FH genes: *LDLR, APOB* and *PCSK9* [38], as well as *LDLRAP1* and *APOE* that have been reported recently to contribute deleterious variants in autosomal recessive [39] and dominant FH [22].

#### SNV discovery

Consistent with previous studies in FH patients from multiple ancestral backgrounds, rare SNVs in *LDLR* contributed the bulk of pathogenic mutations in our cohort. Using the modified ACMG Criteria (see Methods), the AI-based Fabric GEM algorithm that prioritizes variants based on HPO terms [23], and prior functional studies, we identified pathogenic and likely pathogenic variants in 141 individuals from 63 families (**Table 3**). Amongst the 59 families with multiple affected individuals, 59% carried pathogenic variants: twenty-seven were missense, while eight are predicted to induce truncations that trigger nonsense-mediated decay. Amongst singletons, the diagnostic yield was lower, 27 in 108 individuals. This is not surprising, given that elevated LDL cholesterol is common among older individuals (and prominent in Mexico, where hypercholesterolemia affects between 11% and 36% of young men and women [40]). Variants annotated as VUS or benign are listed in **Table S2**.

**Table 3:** Pathogenic Variants Identified in FH Genes (*LDLR*, *APOB*, *PCSK9*, and *LDRAP1*) Within the FH Cohort, Along with Population-Based Frequencies from gnomAD, Functional studies and reported ethnicities of carriers in other studies. Source studies for this data can be found in supplementary references

| Variant | FH cohort (n=300) | GnomAD AF | Functional studies | Ethnicity |
| --- | --- | --- | --- | --- |
| *LDLR*:p.Trp4Ter | 7 het | 0.00000205 | <2% *LDLR* activity [1] | Spain[2] , Mexico[3] |
| *LDLR*:p.Glu101Lys | 2 het | 0.0000294 | 15–30% *LDLR* activity[4] | Spain[5],France[6],UK[4],Mexico[3],Germany[7],Malaysia[8] |
| *LDLR*:p.Cys109Arg | 2 het | 0.00000274 | 15-30% *LDLR* activity[1] | Mexico[3],Germany[9] |
| *LDLR*:p.Glu113Glyfs Ter17 | 12 het | 0.00000137 |  | Mexico[3],[10] |
| *LDLR*:p.Glu140Lys | 2 het and 1 hom | 0.00000342 | 30% *LDLR* activity[11] | South Africa [11], Spain[12],Denmark[13],Italy[14],Korea[15],Mexico[16], Japan[17], |
| *LDLR*:p.Asp168Glu | 6 het | 0.00000137 |  | Turkey [18] |
| *LDLR*:p.Cys173Tyr | 3 het | - |  | Italy[14] |
| *LDLR*:p.Glu228Lys | 5 het | 0.00000549 | 24% on cell surface, 21% binding[19] | China[19],Portugal,Canada[20] [22] |
| *LDLR*:p.Cys289Arg | 2 het | - |  | Mexico[3] |
| *LDLR*:p.Met298Ilefs Ter3 | 9 het | - |  | present study |
| *LDLR*:p.Cys329Tyr | 1 het | 0.00000411 | 31% surface *LDLR*, 31% binding[19] | China [19] |
| *LDLR*:p.Gly343Ser | 1 het | 0.0000356 | 15-30% *LDLR* activity [1] | New Zealand[23],Netherland[24],USA[1],Spain[25] ,Israel[26],Italy[27],Portugal[21] |
| *LDLR*:p.Leu346Alafs Ter12 | 7 het | 0.00000068 |  | Mexico[16] |
| *LDLR*:p.Cys352Tyr | 9 het | 0.00000342 | 15-30% *LDLR* activity[1] | Mexico[28] |
| *LDLR*:p.Cys364Arg | 2 het | 0.00000068 | 15% *LDLR* activity[1] | Mexico[28],New Zealand[29] |
| *LDLR*:p.Cys368Tyr | 6 het | 0.00000274 | 40% reduced binding, 60% reduced uptake[30] | France[6],Spain[25],Mexico[3] |
| *LDLR*:p.Lys393Glu | 3 het | - |  | present study |
| *LDLR*:p.His407Gln | 1 het | 0.00000342 |  | present study |
| *LDLR*:p.Tyr400_Phe402del | 8 het | - | *LDLR* retained in the ER [31] | Spain[5],[32] |
| *LDLR*:p.Thr413Met | 1 het | 0.0000472 | 85% uptake, 80% binding[30] | New Zealand[33],Norway[34],UK[35] |
| *LDLR*:p.Val429Met | 6 het | 0.0000144 | <2% *LDLR* activity[36] | Mexico[16],Italy[14],South Africa[36], South Asia[37] |
| *LDLR*:p.Gln448Ter | 3 het | 0.00000068 |  | Mexico[16],Spain[25] |
| *LDLR*:p.Gly546Val | 2 het and 1 hom | 0.00000068 |  | UK[35],Mexico[16] |
| *LDLR* :p.Asn564His and p.Val800_Leu802del | 2 het | 0.0000137 | 20% *LDLR* activity[38] | Germany[39],Spain[25],French[40],Denmark[38],UK[41],Netherlands[34], Portugal[21],Mexico[16] |
| *LDLR*:p.Ala606Thr | 2 het | 0.00000821 |  | Spain[42] |
| *LDLR*:p.Asp622Gly | 1 het | - |  | Spain[42],Czech[43] |
| *LDLR*:p.Arg633Cys | 1 het | 0.0000328 | Decreased expression, reduced uptake [30] | Spain[25] [30], Netherlands[13],Czech[43], Italy[27],Taiwan[44] |
| *LDLR*:p.Ser648Phe | 1 het | - |  | present study |
| *LDLR*:p.Cys667Phe | 7 het | - |  | UK[45] |
| *LDLR*:p.Cys681Ter | 5 het | 0.00000684 |  | Brazil[46],Norway[47],Lebanon[48],Syria[49],Spain[50],Czech[43],Italy[14],Argentina[51] |
| *LDLR*:p.Pro685Ser | 1 het | - | <75% *LDLR* activity[52] | Portugal[21] |
| *LDLR*:p.Glu714Gln | 4 het | - |  | France[53] |
| *LDLR*:p.Arg744Ter | 1 het | 0.00000753 |  | Singapore[54] |
| *LDLR*:p.Ile792Thr | 2 het | 0.0000267 |  | Spain [42],Italy[27] |

Three variants were recurrent: we found p.Cys352Tyr in four families, p.Glu113GlyfsTer17 in two families and p.Tyr400_Phe402del in two families, with no evidence of direct relatedness, intimating more distant common ancestors. In addition, and consistent with our ancestry data, 39/65 pathogenic *LDLR* SNVs (27 missense, 12 splice/nonsense), 13 have been reported only in patients from Mexico, Italy or the Iberian Peninsula, while five SNVs are novel (**Table 3**).

To probe this observation further, we derived the local ancestry around mutations that were ultra-rare or unreported in gnomAD and asked whether these variants might have arisen in an Amerindian (AMR) background. We found this to be true in numerous instances (**Table 3**). For example, we found *LDLR*:p.Glu113GlyfsTer17 and p.Met298IlefsTer3 primarily in AMR/AMR chromosomes (73% and 86%, respectively). In contrast, *LDLR:*p.Leu346AlafsTer12 was mostly EUR (86%). Similarly, the *PCSK9*:p.Arg218Lys and *LDLRAP1*:c.459+2T>G alleles mapped exclusively in an AMR background. These findings highlight population-specific contributions to these rare variants (**Table 3**; **Fig. 3**) and also suggest that some alleles might have either arisen independently or predate the settlement of the Americas.

**Figure 3:**
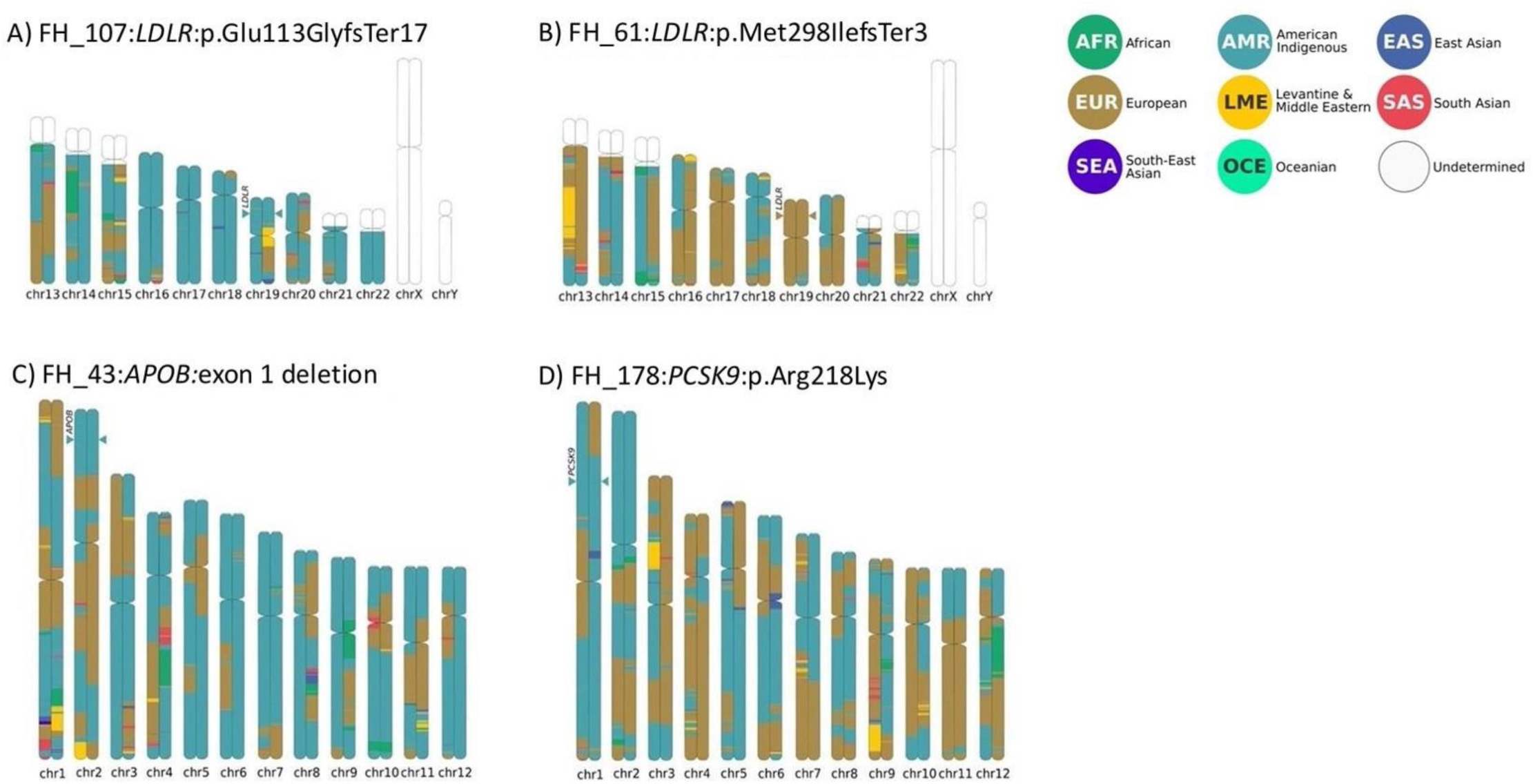
Ancestry ideograms for four samples carrying the variants A) *LDLR*: p.Glu113GlyfsTer17 B)*LDLR*:p.Met298IlefsTer3 C) *APOB*:exon1 deletion, and D) *PCSK9*:p.Arg218Lys. These ideograms illustrate the local ancestry composition at gene loci where pathogenic variants were identified.

We also noted a non-random distribution of mutations across *LDLR* (**Fig. 4**). Six of 27 missense variants map to exon 4, followed by further clustering in exons 6, 8, 9 and 14. Exon 4 encodes the ligand binding domain and has been found to harbor the highest number of pathogenic mutations in FH patients, even after adjusting for size of exons (**Fig. 4**) [41].

**Figure 4.**
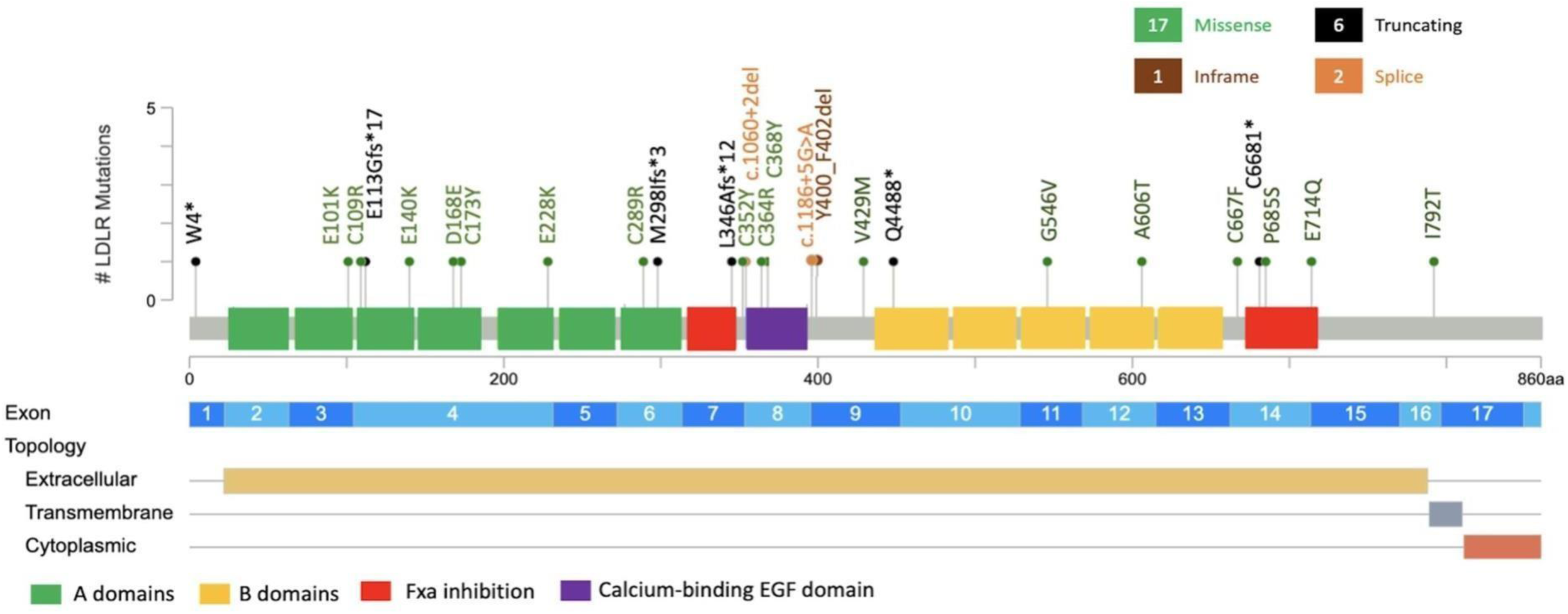
Lollipop plot illustrating the clustering of 40 pathogenic or likely pathogenic variants located within the exonic and intronic regions of the *LDLR* gene.

*APOB* variants were more challenging to interpret; the locus harbors numerous rare and ultra-rare variants of unknown functional effect, with scant evidence to enable inclusion or rejection from a role in FH. Consistent with prior studies that suggested *APOB* to be a rare cause of FH in Mexico [42], we found a single SNV (p.Tyr3560Cys) that segregated in the three affected members of one family. This allele has been implicated in FH in patients from Portugal [43] and Australia [44], and was absent from our internal control cohort (n=1,500 chromosomes) All remaining candidate variants (**Table S2**) bore either inconclusive genetic evidence and/or lacked functional insights. Notably, we did not find any instances of the R3500W variant, the most frequent *APOB* mutation in Northern European populations [45]. Similarly, the less frequent but recurrent Northern European alleles 3500Q and R3531C [45] were likewise absent.

Finally, we found deleterious SNVs in each of *APOE* and *LDLRAP1*. For *APOE*, we detected a three-amino acid deletion (p.Leu167del), shown previously to cause adult Dominant Hypercholesterolemia [22] and FH [46]. In *LDLRAP1*, the only recessive locus in this series, we found a single individual bearing a homozygous splice variant (c.459+2T>G) reported previously in patients from Mexico[47], unrelated to the individual in our study.

### CNV analysis

Previous studies have supported that structural variation accounts for a considerable percentage of FH cases. For example, copy number variations (CNVs) in *LDLR* have been shown in as much 10% of FH cases [48]. Here, using a combination of exome depth coverage and orthogonal validations by Taqman PCR assays, we found two CNVs in LDLR (**Table 3 and Figure S1**): a deletion of exons 13-14 that segregated with the disease in all three patients from family 107 and a deletion of exons 1-15 in the sole affected individual of family 82.

In contrast to *LDLR*, CNVs have not been reported for *APOB*. In our cohort, however, we found a deletion of exon 1 of *APOB* in four unrelated individuals (families 165, 196, 33 and 84). The variant is likely pathogenic since it deletes the start Methionine and induces a frameshift mutation and premature termination. Notably, the patient from family 165 was also heterozygous for the known pathogenic *LDLR* variant p.Lys393Glu, raising the possibility that the two variants may act synergistically to contribute to FH. We failed to find the *APOB* deletion in 741 internal, ancestry-adjusted normolipidemic controls as well as in the gnomAD and the UKB databases.

Finally, we identified an individual with a duplication of *PCSK9* described previously in two unrelated Canadian FH patients[49].

All observed CNVs were confirmed by custom Taqman RT-PCR assays, which were also used for segregation analysis. In addition, for the families bearing the *LDLR* exon 13-14 CNV and the *PCSK9* duplication, we had sufficient DNA to perform Illumina short-read whole-genome sequencing and Oxford Nanopore long-read sequencing. In addition to confirming these lesions with both technologies, we also mapped all four breakpoints. For *LDLR*, both the right and left breakpoints mapped within *Alu*SP sequences in introns 12 and 14 respectively that shared 86% sequence similarity and were oriented in the same direction, supporting an *Alu*-*Alu* recombination mechanism (**Figure S1**). We found the same genomic organization in *PCSK9*: breakpoint analysis identified *Alu*SN sequences at the breakpoint junctions. These elements also exhibited 85% similarity and maintained the same orientation (**Figure S1**).

### Phenocopy rate

Amongst the 59 families with multiple members available to this study, we identified 24 patients who did not inherit the family mutation but were diagnosed with FH because of their lipid profile and family history. In Family 7, for example, we identified four patients who carry the pathogenic *LDLR* p.C352Y variant, along with one first-degree relative who tested negative for the mutation yet had been considered affected by virtue of family history, high LDL cholesterol (189) and low HDL cholesterol (29). Considering our entire cohort, but restricting our analysis to families with 2+ affected individuals, we found a maximal phenocopy rate of 12.5% (24/192 individuals), likely driven by the fact that FH is clinically indistinguishable from sporadic hyperlipidemia, a common disease.

### Overlap with primary dyslipidemias and lipodystrophies

Variants in *ABCG5*, *ABCG8*, and *LPA* have been causally associated with sitosterolemia, a condition that shares symptoms with FH, including elevated cholesterol levels, xanthomas, and increased cardiovascular risk [50] while variants in *ABCA1* can cause hypoalphalipoproteinemia, characterized by low plasma HDL levels [51]. However, the two conditions differ in terms of pathophysiology: sitosterolemia is characterized by the accumulation of dietary plant sterols in the blood and tissues due to defective sterol transporters, whereas FH is marked by elevated levels of LDL cholesterol due to impaired LDL clearance. Furthermore, sitosterolemia is typically found as an autosomal recessive trait.

None of our patients carried homozygous or compound heterozygous variants in *ABCA1*, *ABCG5*, or *ABCG8*. In one patient we found two variants in *ABCG5* (p.R105L and p.A98G); the first variant is absent from all databases, while the second has been reported once in an FH patient in the ClinVar database, is conserved across species, but is predicted to be tolerated, thereby receiving a preliminary designation as a VUS. Similarly, we found two VUSs in *LIPA* (**Table S3**).

However, we did discover two pathogenic variants, one in *PLIN1* and one in *CREB3L3*, each reported previously in dominant familial partial lipodystrophy (FPL) [52] and hypertriglyceridemia [53] (**Figure S2**). Common signs with FH include high levels of triglycerides and dyslipidemia in patients with FPL while patients with hypertriglyceridemia have high levels of total cholesterol, triglycerides and low levels of HDL [53]. Although we cannot exclude the possibility that these are patients misdiagnosed with FH, we consider that possibility unlikely; typical FPL patients exhibit a striking loss of adipose tissue in their limbs and hips, which we did not observe. Instead, we favor the possibility that these cases represent phenotypic expansion for *PLIN1* and *CREB3L3*.

### Diagnostic yield

Focusing on families with multiple affected members (and as such more likely to capture a true FH diagnosis), we identified pathogenic variants in 125/192 patients across 40 of 59 families, resulting in a diagnostic yield of 65%. Unsurprisingly, the diagnostic yield in singleton cases was lower at 32% (35/108 cases), further underscoring the importance of family history as a criterion for FH diagnosis. **Fig. 5** illustrates the distribution of allele types in our cohort.

**Figure 5:**
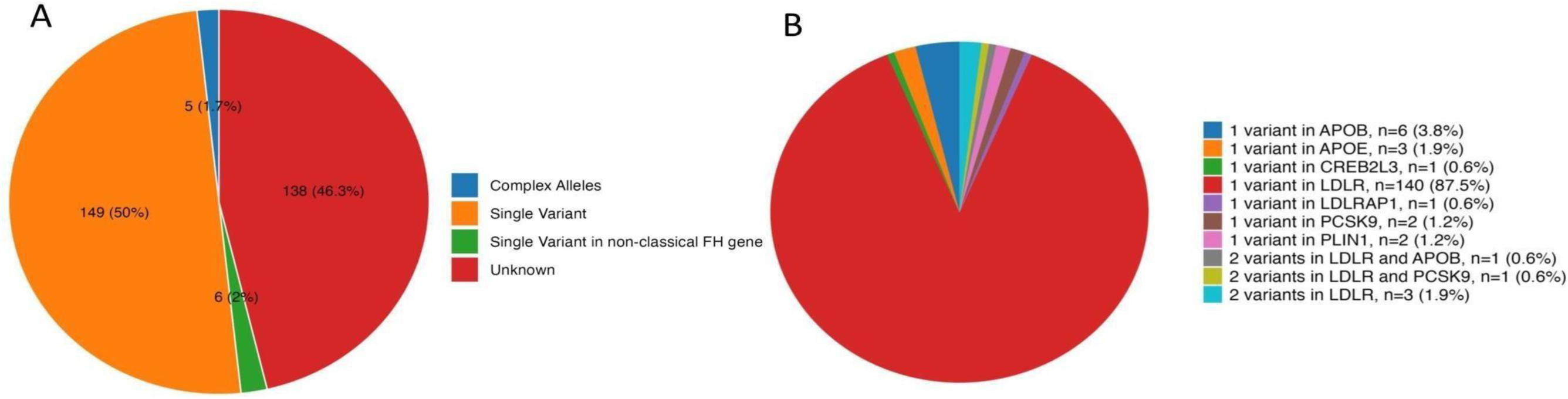
A) Restricting the analysis to only pathogenic or likely pathogenic variants, distribution of allele types in our cohort: single variant, complex alleles, non-classical gene, and unknown. B) Distribution of variants found in the 3 major FH genes as well as in FH phenocopy genes.

### Dissection of the remaining missing variation

We turned to the question of genetic causality in the 35% of patients who lacked a molecular diagnosis, which even under the assumption of a 12.5% phenocopy rate, remains substantial. We explored two possibilities: a) that the remainder of the genetic burden for FH is driven by common variants; and b) that some of our mutation-negative patients might harbor rare, high liability alleles in hitherto unknown genes.

### Polygenic Risk Score Analysis

To answer the first question, we computed the polygenic risk score for high LDL across our entire cohort (irrespective of their genetic burden in the known FH^+^ genes) and compared that to the PRS distribution of a control cohort.

Our LDL PRS model predicted significantly different risk scores between the FH cohort and controls (**Fig. 6**). Among the patients with a definitive molecular diagnosis, 10.6% mapped above the 95^th^ centile of LDL PRS distribution, compared to ancestry-adjusted controls (3.0%; **Fig. 6A, B**). The enrichment was even more pronounced among the FH patients without a molecular diagnosis, 19.7% of whom scored above the 95th centile of risk. By using a Exact Fisher’s Exact Tail with Holm-Bonferroni correction, we observe the difference in the 95^th^ centile tails -to be significant when compared to either controls (p-value < 0.001) or non-carriers of pathogenic variants (p-value =0.03). Furthermore, the differences between the average scores between groups were also significant when applying a Welch’s t-test: when comparing cases vs controls (p-value < 0.001) and carriers vs non-carriers (p-value < 0.001). By applying a linear regression from ancestry-adjusted scores to cumulative LDL years, we observed that individuals at the tail of the distribution carried the same risk as individuals with high liability heterozygous pathogenic alleles, suggesting that some patients do not necessarily carry rare, high liability alleles but distributed additive risk from common variants (**Fig. 6C**). Finally, we measured the risk scores using over 4000 models available on the PGSCatalog (www.pgscatalog.org) and after applying a Benhamin-Hochberg correction, a total of 17 models showed a significant p-value from which five are LDL models, and 14 are cholesterol-related (**Figure S3**).

**Figure 6:**
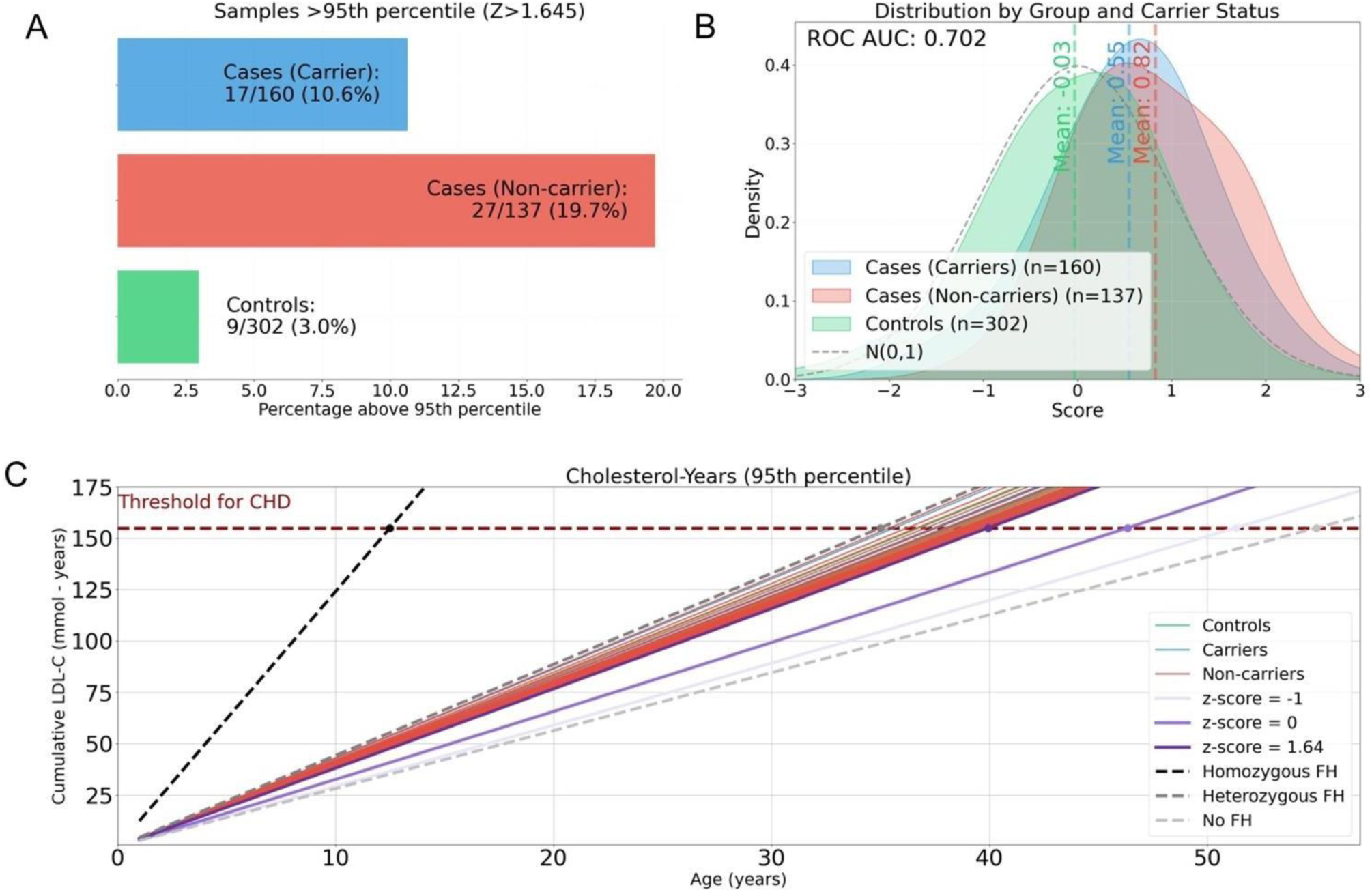
A) LDL PRS distribution for controls and cases (carriers and non-carriers) B) Percentage of samples above the 95th percentile of a Normal distribution C) Cumulative incidence of LDL given the age of the patient

#### Variants in novel genes encoding cholesterol pathway components

Finally, we asked whether some of the remaining affected FH individuals might carry rare, high liability variants in hitherto unknown FH genes. To test this idea, we focused on the 47 genes known to encode components of cholesterol biosynthesis and transport and an additional 55 genes that encode 1^st^-order interactors (total 102 genes, see Methods).

We first filtered for loci that fulfill the following three criteria: a) they carry non-synonymous, predicted deleterious alleles with a population frequency below the carrier frequency of FH (1:200 [54]) in public databases such as UKB and gnomAD; b) such alleles ought to be absent from our internal, normolepidemic, ancestry-adjusted controls; and c) they either show evidence of segregation within our mutation-negative families or show recurrent alleles in at least two undiagnosed, unrelated individuals.

We did not detect any point mutations or small indels that, under our assumptions, offered compelling evidence of locus involvement in FH. Although this observation might reflect over-filtering, it also reflects the fact that FH is a typically dominant, relatively common disease with a high phenocopy rate and, as such remains prone to false positives.

In contrast, we identified a CNV that intrigued us. Specifically, two unrelated (pi-hat=0) individuals in our cohort carry a whole-gene duplication of *PDZK1* (**Fig. 7A**). PDZK1 encodes a scaffolding protein that interacts with membrane proteins, including the high-density lipoprotein (HDL) scavenger receptor class B type I (SR-BI) [55]. Typing our control set, we were encouraged to see the variant absent from 741 normolipidemic individuals.

**Figure 7:**
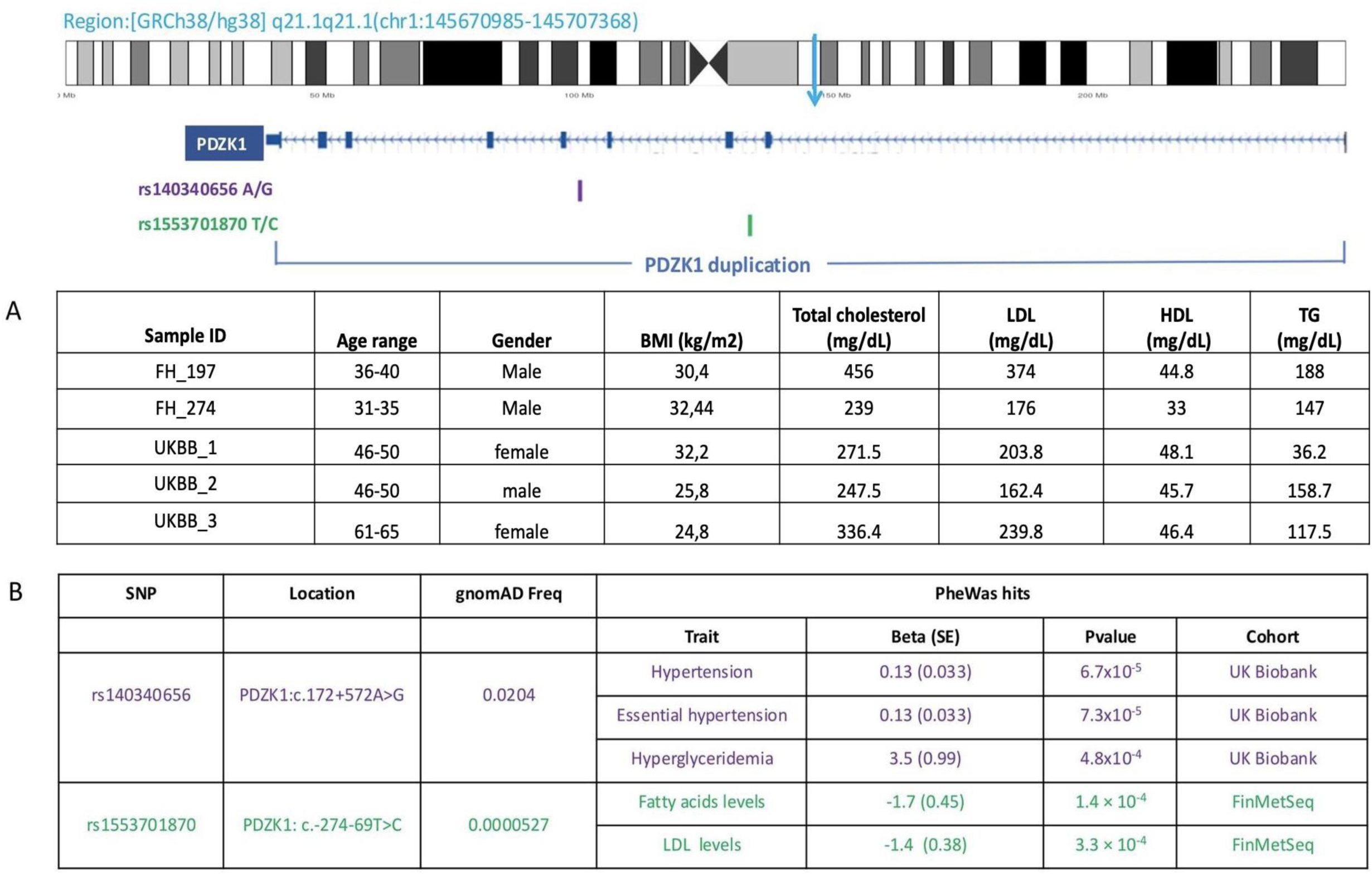
Location of the duplication in *PDZK1* gene. A) demographic/clinical characteristics of the 5 carriers identified in FH cohort and UK Biobank. B) PheWAS results for two intronic variants (rs140340656 and rs1553701870) in PDZK1. The PheWAS results were obtained from the pheWeb database including data from studies in the UK Biobank and FinMet cohorts.

Given these findings, we turned to the UKB, where we analyzed array data from 498,200 individuals. We identified three carriers of the *PDZK1* duplication; in each case, their LDL and total cholesterol levels mapped to the top quartile of the entire cohort for LDL and total cholesterol, with two individual recording LDL-C >190 and the third individual recording elevated LDL-C (163). (**Fig. 7A**).

### PheWas studies

To investigate further the possible involvement of *PDZK1* variation in FH, we mined the PheWeb database for relevant phenotypes that include a clinical diagnosis of dyslipidemia, hypertension, or cardiovascular disease, as well as direct lipid measurements, all phenotyped in large cohorts, such as the UK Biobank, the Michigan Genomics Initiative, and FinMetSeq [56].

We found two intronic variants and one intergenic variant 41kb upstream of *PDZK1* to be associated with hypertension and lipid levels (**Fig. 7B**). Specifically, the intronic variant c.793+572T>C (rs140340656) showed association with an elevated risk for hypertension (Beta = 0.13, SE = 0.033, p = 6.7 × 10⁻⁵), essential hypertension (Beta = 0.13, SE = 0.033, p = 7.3 × 10⁻⁵), and hypertriglyceridemia (Beta = 3.5, SE = 0.99, p = 4.8 × 10⁻⁴) in the UK Biobank. The second intronic variant, c.211-69A>G (rs1553701870), exhibits a protective effect in FinMetSeq, with significantly lower levels of saturated fatty acids (Beta = −1.7, SE = 0.45, p = 1.4 × 10⁻⁴) and LDL cholesterol (Beta = −1.4, SE = 0.38, p = 3.3 × 10⁻⁴). Finally, the intergenic variant rs1649823291 was associated with reduced levels of APOB abundance (Beta = −1.1, SE = 0.33, p = 5.6 × 10⁻⁴), which has been shown to be protective of cardiovascular disease and mortality secondary to heart attack and stroke [57].

## Discussion

Here we have presented our study of a 300-strong cohort from the Mexican FH registry. Consistent with previous findings across populations, we attribute >80% of FH cases with a definitive molecular diagnosis to dominant mutations in *LDLR* [58]. The most common mutation was an insertion in exon 4 (p.Glu113GlyfsTer17), reported previously in Mexican FH patients [59]. Likewise, another three variants (p.Cys352Tyr, p.Leu346AlafsTer12, p.Cys289Arg) have only been reported in the Mexican population [42][60][61]. More broadly, almost a third of the *LDLR* mutations in our cohort have only been seen in Spanish, Portuguese, and Italian populations, but not in Northern Europeans (despite the supernumerary studies in the latter). In one family, we also found the p.Cys681Ter allele, known to account for 60% of FH cases in Lebanon [62], but also reported in other Latin American cohorts [63]. The enrichment of alleles from Mediterranean Europe and the Middle East reflects the distinct genetic background of these geographic regions and their contribution to the genetic architecture of Latin America [63]. Of note, six variants (p.Met298IlefsTer3, p.Lys393Glu, p.His407Gln, p.Ser648Phe, c.1060+2del, deletion of exon1-15) are novel and might represent variants arisen in a Native American background. Discovery of such alleles remains critical, both for improving diagnostic capabilities in admixed individuals and for informing the biology of the disorder.

*APOB* and *PCSK9* mutations were rare in our cohort. Of note, the most frequent FH-causing *APOB* mutation in “Europeans”, p.Arg3500Gln [64], was absent, further underscoring the fact that “Europeans” represent a sometimes simplistic label with limited utility in interpreting genetic variation in admixed Latin American populations. Moreover, among the rare variants we found in *APOB*, only two offer sufficiently compelling evidence for pathogenicity. The first is a p.Tyr3560Cys variant, reported previously in FH patients of Portuguese [65] and Dutch ancestry [44]. The second is a novel, heterozygous deletion of exon 1; we do not know whether the discovery of this allele reflects a variant arisen in a Native American background, or that single-exon CNVs are often disregarded or not detected in exome-based diagnostic tests [66].

Extending our analysis to non-classical FH genes, we identified several pathogenic variants, starting with a p.Leu167del variant in *APOE*. This variant, reported previously in FH patients of Italian [22] and Spanish [67] descent, disrupts the leucine zipper near the 4th α-helical structure of *APOE*, a region that mediates its interaction with *LDLR*. The mutation increases the binding affinity of *APOE* to *LDLR*, preventing receptor transport back to the hepatocyte surface to clear cholesterol [67]. Carriers of the p.Leu167del mutation exhibit a better response to lipid lowering therapy than *LDLR* carriers[68] further motivating the addition of *APOE* in FH genetic panels. Extending that idea, we have also found *bona fide* pathogenic mutations in *PLIN1* and *CREB3L3*, each found previously in familial partial lipodystrophy (FPL). However, given that our patients do not exhibit the adipose deposits that hallmark FPL, we once again suggest that a more “lipodystrophy” centered approach to analyzing panels, exomes and genomes for FH might serve patients and their families better.

In multiplex families, our diagnostic yield approaches 70%, at the upper bound of reported work [69] and significantly higher than cross-sectional population-based studies, where some studies have reported a mutational yield in the single digits [70]. In our cohort, even in singletons, our diagnostic yield exceeded 30%. We attribute this diagnostic yield to the stringent application of the Dutch diagnostic criteria for FH; to the expansion of genes interrogated to include both loci that cause other clinical forms of dyslipidemia and the remaining components of the cholesterol pathway; and the accurate detection of CNVs, which contribute 5-10% to the mutational burden but are often undetected in targeted sequencing clinical tests. We also note that application of appropriate PRS analysis has revealed two points that merit attention. First, we observed that, even in the presence of a rare, high liability allele in a known FH gene, a significant fraction of our patients maps to the 95^th^ or 99^th^ centile of PRS-derived risk, suggesting that common alleles likely exert an effect on rare, in principle penetrant mutations. Longitudinal studies on disease onset, progression and cardiovascular outcomes, as well as response to a variety of medications, will be necessary to understand the effect of such variation on the phenotype. In addition, we also found that the PRS signal was significantly more pronounced in *bona fide* FH patients bereft of high liability alleles, arguing that individuals will manifest FH because of their common allele burden and not rare, peri-Mendelian variants. In that context, we support the development of clinical PRS tools and suggest that, for some disorders such as FH, they might have diagnostic and clinical utility.

Finally, we draw attention to the fact that most clinical genetic tests for FH (and other disorders) focus on single nucleotide variants and small indels, even though CNVs can account for as much as 10% of the genetic burden [48]. Our findings further underscore the importance of structural variants in FH. The continued development of robust CNV analysis and validation tools, both through improved exome methods and by transitioning to whole-genome sequencing will be important to support molecular diagnosis and new gene discovery for FH and other disorders.

The detection of clinically-relevant duplications can be challenging, yet carry significant ramifications. For example, one of our patients was a carrier of a whole-gene duplication of *PCSK9*. This finding has treatment implications, as statin-induced upregulation of *PCSK9* has been suggested to be less effective due to the presence of an additional *PCSK9* copy [49]. Second, duplication analysis was a catalyst for the discovery of a novel candidate FH locus via the identification, and subsequent molecular confirmation of a duplication in *PDZK1* in two unrelated patients. We recapitulated this finding in the UKB, where we found the same lesion in three individuals, all of whom showed elevated lipid profiles. The absence of any phenotypic signal for loss-of-function events at this locus suggests that dosage sensitivity is the most likely contributory mechanism to FH **(Fig. S5).** Consistent with this notion, we identified two intergenic and one intragenic SNP that show association with hypertension and lipid levels. We are particularly intrigued by the fact that one of the SNPs (rs1553701870) likely exerts a protective effect. Based on population-wide PheWAS of UKB participants with loss-of function *PDZK1* mutations and a deeper analysis of ∼600 individuals with gene-disruptive deletions, we suggest that loss-of function at this locus is not likely to impart obvious pathology in humans. Indeed, heterozygous knockout mice are asymptomatic, while homozygous null *PDZK1* mice likewise exhibit normal liver and renal physiology, with albeit increased HDL abundance [55]. Together, the human and murine data suggest that suppression or ablation of *PDZK1* might be of therapeutic benefit and offer the development of a class of drugs orthogonal to the current set of offerings such as statins and *PCSK9* inhibitors. We are intrigued by the recent association of *PDZK1* with gout [71], a phenotype strongly associated with dyslipidemia, most notably in a meta-analysis of 514,000 individuals from the Korean National Health Insurance Service-Health Screening Cohort [72]. We speculate that the renal, not the liver role(s) of *PDZK1* might be most relevant to its therapeutic potential and merit further investigation. More broadly, this study highlights the persistent and urgent need to study well-phenotyped cohorts across the globe and to combine iterative learnings that can inform and ultimately improve patient management and public health policy.

## Data Availability

All data produced in the present study are available upon reasonable request to the authors

## Declarations

### Ethics approval and consent to participate

This research protocol has been approved by the Institutional Research and Ethics Committees (IRB) of the National Institute of Health Sciences and Nutrition “Salvador Zubirán”, in Mexico City, under reference number 2571. The sample identifiers used in this study are known only to the research team and do not correspond to any identifiers recognized by individuals outside the study, including hospital staff or participants themselves

### Availability of data and materials

Individual-level data used in this research were collected in a healthcare setting and are subject to controlled access for privacy reasons. The ethics committee and/or informed consent does not allow for public availability.

This research has been conducted using the UK Biobank Resource under Application Number 89006

### Authors’ contributions

Conceptualization, N.K, C.D.B, C.A.A.-S; Genomic analysis: M.T.M.L, M.A.F; Bioinformatics: N.M, C.D.Q, A.L.P, S.M-G, D.M.M,M.B,D.B,P.S.F; Supervision: N.K, C.D.B, C.A.A.-S, A.J.M, A.G.I,F.M.D.L.V,J.G,J.W,B.M; Clinical data curation: FH Mexican Registry Group, A.J.M, C.A.A.-S, R.M, G.A.G-R,R.Z,D.E-L,T.T-L; Manuscript writing: N.K, N.M; Revision of the manuscript: all authors; Funding: FH Mexican Registry Group

## Acknowledgements.

The authors thank all the patients and their families for participating in this study. Authors express their gratitude to Amgen, for the provision of an unrestricted grant for a physician initiated project. The sponsor did not have access to the clinical data, and had no participation, nor influence in the preparation of the manuscript. Our gratitude also, to the investigators and patients who have agreed to participate in the Mexican FH Registry.

