## Supplementary Material for "Analysis of a deeply-phenotyped familial hypercholesterolemia cohort from Mexico shows a role for both rare and common alleles across known dyslipidemia genes and reveals structural variation in a novel locus"

**Supplementary Figure 1:** (A) Schematic representation of the breakpoints and Alu sequences in introns 12 and 14 of *LDLR*. Exons are depicted as vertical lines, while the horizontal line represents introns. Colored triangles indicate the Alu repeats at the breakpoints, with their orientation denoted by the direction of the triangles. The Alu repeats at both sides of the deletion share high sequence similarity (84%) and are aligned in the same orientation, consistent with a mechanism of Alu-Alu recombination (B) Schematic representation of the breakpoints and AluSN repeats flanking the duplication. These repeats exhibit high homology, with 85% sequence similarity, and are also in the same orientation, supporting an Alu-Alu recombination mechanism.

**Supplementary Figure 2:** Pedigrees of the families with A) *APOE* p.Leu167del variant B) *PLIN1* p.Arg93Ter variant C) *CREB3L3* p.Leu461Met variant

**Supplementary Figure 3:** Number of significant genetic models from the PGSCatalog ([www.pgscatalog.org](http://www.pgscatalog.org)) after applying a Benjamini-Hochberg correction. The x-axis represents the traits of the models, and the y-axis shows the number of significant models.

**Table S1**: List of genes participating in the cholesterol pathway used in this study (KEGG pathway: hsa048979) along with their interactors acquired from the Intact database.

**Table S2:** VUS and benign variants in FH genes (*LDLR,APOB,PCSK9*) identified in FH Cohort, along with Population-Based Frequencies from gnomAD and in-house controls.

**Table S3:** Variants in FH phenocopies genes (ABCG5,ABCG8,ABCA1 and LPL) identified in FH Cohort, along with Population-Based Frequencies from gnomAD and ClinVar annotation.

**Supplementary references**

**Supplementary Figure 1**

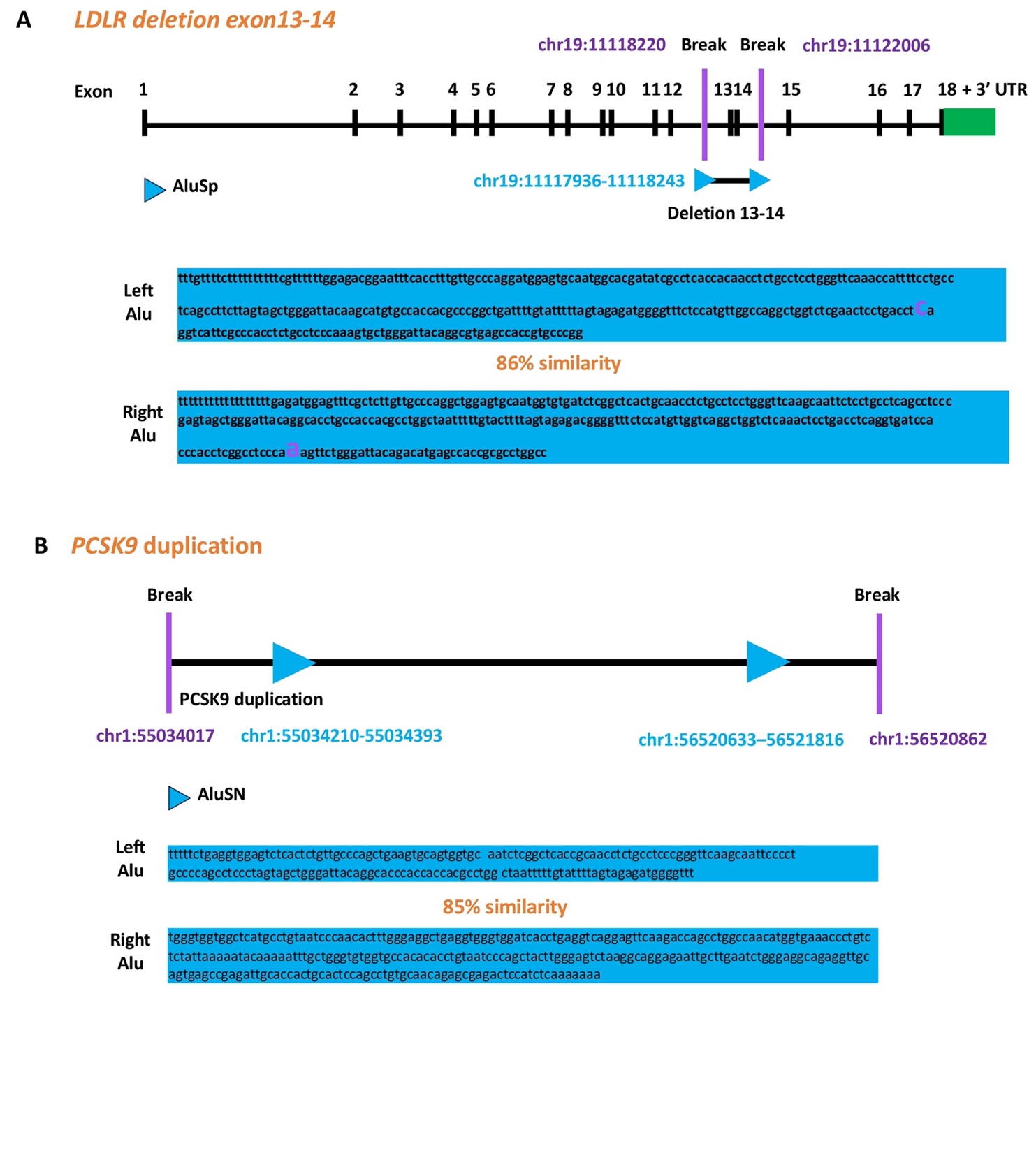

**Supplementary Figure 2**

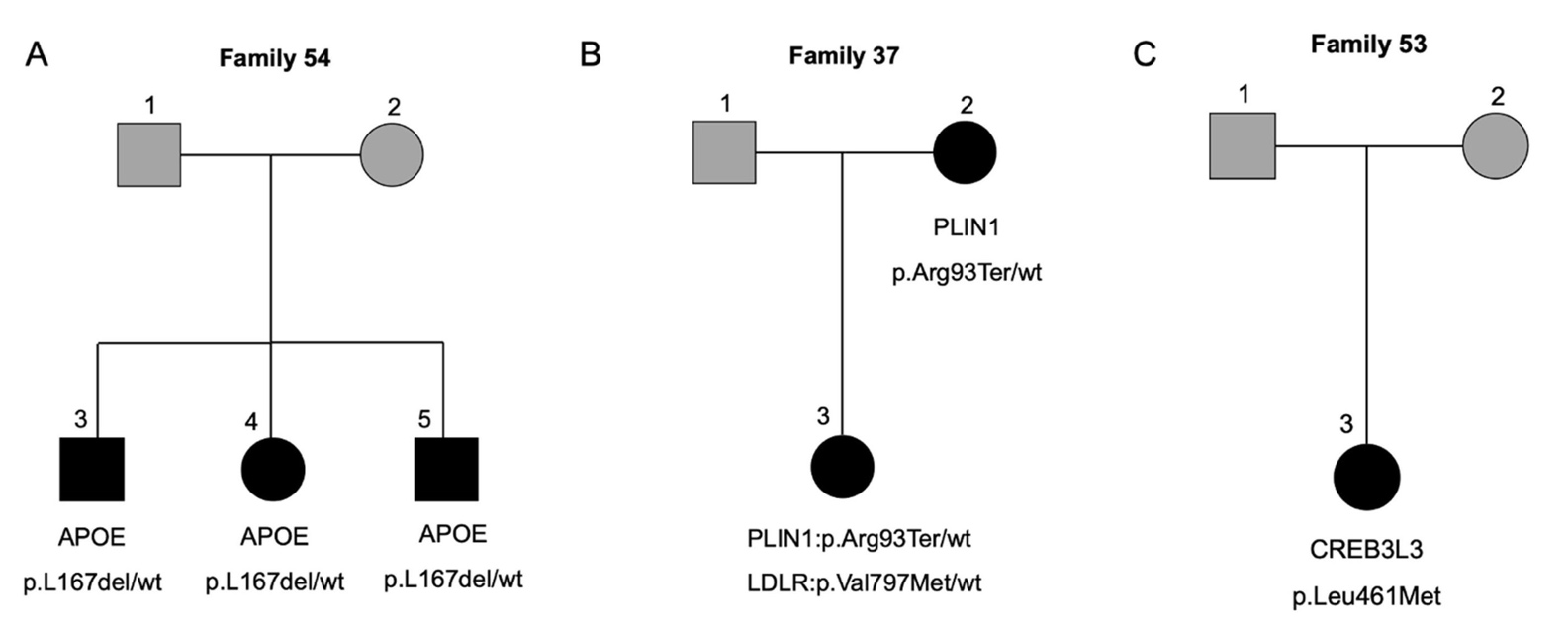

**Supplementary Figure 4**

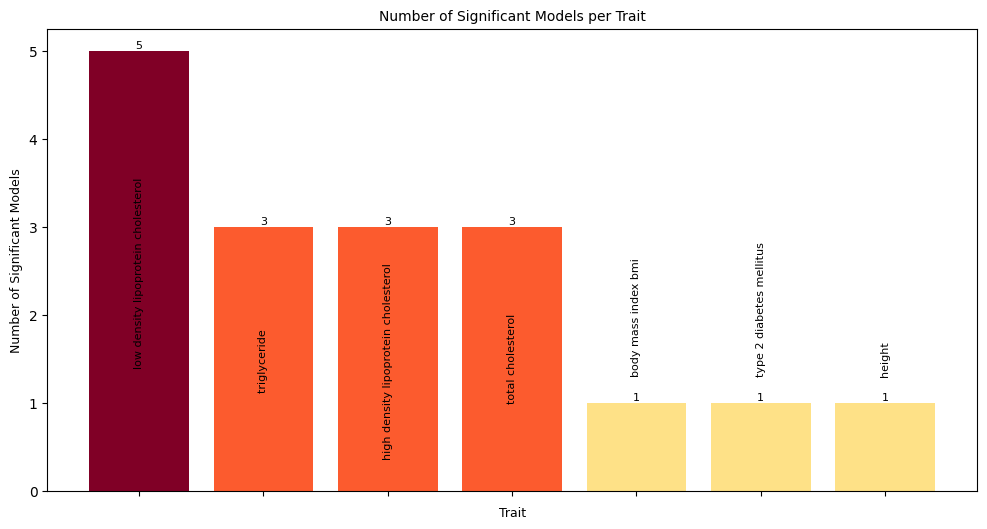

**Table S1**

| **Chromosome** | **Start** | **End** | **Gene** | **Pathway** |
| --- | --- | --- | --- | --- |
| chr9 | 104781006 | 104928155 | *ABCA1* | cholesterol pathway |
| chr15 | 60347151 | 60397986 | *ANXA2* | cholesterol pathway |
| chr11 | 116835751 | 116837622 | *APOA1* | cholesterol pathway |
| chr1 | 161222292 | 161223628 | *APOA2* | cholesterol pathway |
| chr19 | 44946051 | 44949565 | *APOC2* | cholesterol pathway |
| chr12 | 124776856 | 124863864 | *SCARB1* | cholesterol pathway |
| chr16 | 56961950 | 56983845 | *CETP* | cholesterol pathway |
| chr16 | 67939750 | 67944120 | *LCAT* | cholesterol pathway |
| chr8 | 19939253 | 19967259 | *LPL* | cholesterol pathway |
| chr18 | 49562057 | 49599185 | *LIPG* | cholesterol pathway |
| chr20 | 45898622 | 45912155 | *PLTP* | cholesterol pathway |
| chr15 | 58431991 | 58569844 | *LIPC* | cholesterol pathway |
| chr6 | 160531482 | 160664275 | *LPA* | cholesterol pathway |
| chr7 | 80638642 | 80679274 | *CD36* | cholesterol pathway |
| chr3 | 172630249 | 172711067 | *NCEH1* | cholesterol pathway |
| chr12 | 53103486 | 53124535 | *SOAT2* | cholesterol pathway |
| chr1 | 179293797 | 179358680 | *SOAT1* | cholesterol pathway |
| chr19 | 44914608 | 44919346 | *APOC1* | cholesterol pathway |
| chr17 | 66212033 | 66229415 | *APOH* | cholesterol pathway |
| chr1 | 62597520 | 62606313 | *ANGPTL3* | cholesterol pathway |
| chr19 | 8364155 | 8374370 | *ANGPTL4* | cholesterol pathway |
| chr19 | 11239619 | 11241943 | *ANGPTL8* | cholesterol pathway |
| chr1 | 109309575 | 109397918 | *SORT1* | cholesterol pathway |
| chr6 | 16129086 | 16148248 | *MYLIP* | cholesterol pathway |
| chr12 | 57128483 | 57213361 | *LRP1* | cholesterol pathway |
| chr2 | 169127109 | 169362534 | *LRP2* | cholesterol pathway |
| chr17 | 39637144 | 39664201 | *STARD3* | cholesterol pathway |
| chr5 | 133971871 | 134004975 | *VDAC1* | cholesterol pathway |
| chr10 | 75210796 | 75231448 | *VDAC2* | cholesterol pathway |
| chr8 | 42391880 | 42405937 | *VDAC3* | cholesterol pathway |
| chr22 | 43151559 | 43163242 | *TSPO* | cholesterol pathway |
| chr2 | 218782147 | 218815293 | *CYP27A1* | cholesterol pathway |
| chr18 | 9914016 | 9960021 | *VAPA* | cholesterol pathway |
| chr20 | 58389229 | 58451101 | *VAPB* | cholesterol pathway |
| chr18 | 23531442 | 23586506 | *NPC1* | cholesterol pathway |
| chr11 | 3087107 | 3165310 | *OSBPL5* | cholesterol pathway |
| chr14 | 74479940 | 74493305 | *NPC2* | cholesterol pathway |
| chr8 | 38142700 | 38150952 | *STAR* | cholesterol pathway |
| chr14 | 24305187 | 24307977 | *CIDEB* | cholesterol pathway |
| chr8 | 58490178 | 58500163 | *CYP7A1* | cholesterol pathway |
| chr2 | 168920781 | 169031324 | *ABCB11* | cholesterol pathway |
| chr11 | 116829907 | 116833072 | *APOC3* | cholesterol pathway |
| chr11 | 116820700 | 116823304 | *APOA4* | cholesterol pathway |
| chr2 | 43812472 | 43838839 | *ABCG5* | cholesterol pathway |
| chr2 | 43838971 | 43882988 | *ABCG8* | cholesterol pathway |
| chr10 | 89213572 | 89251775 | *LIPA* | cholesterol pathway |
| chr4 | 3503612 | 3532422 | *LRPAP1* | cholesterol pathway |
| chr19 | 44946051 | 44949565 | *APOC2* | interactor |
| chr14 | 74479940 | 74493305 | *NPC2* | interactor |
| chr18 | 23531442 | 23586506 | *NPC1* | interactor |
| chr1 | 179293797 | 179358680 | *SOAT1* | interactor |
| chr1 | 109309575 | 109397918 | *SORT1* | interactor |
| chr21 | 25880550 | 26170770 | *APP* | interactor |
| chr10 | 52765380 | 52772784 | *MBL2* | interactor |
| chr5 | 75377775 | 75511629 | *CERT1* | interactor |
| chr1 | 159587826 | 159588865 | *APCS* | interactor |
| chr15 | 74782080 | 74803197 | *CSK* | interactor |
| chr20 | 41137543 | 41177626 | *PLCG1* | interactor |
| chr22 | 30240453 | 30246759 | *LIF* | interactor |
| chr11 | 119185475 | 119190213 | *NHERF4* | interactor |
| chr12 | 14612632 | 14696599 | *GUCY2C* | interactor |
| chr17 | 58301231 | 58328795 | *TSPOAP1* | interactor |
| chr1 | 145670985 | 145707368 | *PDZK1* | interactor |
| chr5 | 157477304 | 157575775 | *ADAM19* | interactor |
| chr11 | 116789367 | 116791879 | *APOA5* | interactor |
| chr5 | 38474668 | 38556646 | *LIFR* | interactor |
| chr3 | 124761948 | 124887365 | *ITGB5* | interactor |
| chr11 | 1752755 | 1763927 | *CTSD* | interactor |
| chr5 | 55935095 | 55994963 | *IL6ST* | interactor |
| chr8 | 143213218 | 143217170 | *GPIHBP1* | interactor |
| chr15 | 39581079 | 39599466 | *THBS1* | interactor |
| chr1 | 115285917 | 115338249 | *NGF* | interactor |
| chr1 | 156860865 | 156881850 | *NTRK1* | interactor |
| chr16 | 30896614 | 30903547 | *CTF1* | interactor |
| chr11 | 58622665 | 58625733 | *CNTF* | interactor |
| chr10 | 106573663 | 107164706 | *SORCS1* | interactor |
| chr8 | 132866958 | 133134899 | *TG* | interactor |
| chr16 | 23463542 | 23510494 | *GGA2* | interactor |
| chr6 | 160702244 | 160754097 | *PLG* | interactor |
| chr22 | 30262829 | 30266851 | *OSM* | interactor |
| chr12 | 85874295 | 85882992 | *NTS* | interactor |
| chr2 | 215360865 | 215436068 | *FN1* | interactor |
| chr22 | 19846146 | 19854874 | *RTL10* | interactor |
| chr4 | 76158737 | 76213824 | *SCARB2* | interactor |
| chrX | 101397803 | 101407925 | *GLA* | interactor |
| chr18 | 24162052 | 24397824 | *OSBPL1A* | interactor |
| chr9 | 134880810 | 134887523 | *FCN2* | interactor |
| chr17 | 44345302 | 44353106 | *GRN* | interactor |
| chr9 | 87497867 | 87708634 | *DAPK1* | interactor |
| chr22 | 37608834 | 37633564 | *GGA1* | interactor |
| chr15 | 87859751 | 88256739 | *NTRK3* | interactor |
| chr10 | 32900318 | 32958230 | *ITGB1* | interactor |
| chr11 | 6395125 | 6419094 | *APBB1* | interactor |
| chr10 | 97737128 | 97760895 | *ZFYVE27* | interactor |
| chr16 | 72054505 | 72061055 | *HP* | interactor |
| chr8 | 11842524 | 11868087 | *CTSB* | interactor |
| chr11 | 27654893 | 27700455 | *BDNF* | interactor |
| chr7 | 22727200 | 22731998 | *IL6* | interactor |
| chr20 | 45469753 | 45481532 | *WFDC2* | interactor |
| chr16 | 2026902 | 2039026 | *NHERF2* | interactor |
| chr9 | 84669729 | 85027054 | *NTRK2* | interactor |
| chr6 | 33572552 | 33580276 | *BAK1* | interactor |

**Table S2**

| **Gene** | **carriers FH cohort**  **(n=300)** | **carriers controls**  **(n=741)** | **GnomAD exomes Freq** |
| --- | --- | --- | --- |
| **VUS** |  |  |  |
| *LDLR*:p.His211Tyr | 1 het | 3 het | 0.00000274 |
| *LDLR*:p.Glu28Lys | 5 het | 5 het | 0.0000616 |
| *LDLR*:c.2312-12T>G | 1 het |  | - |
| *LDLR*:c.190+12T>A | 1 het |  | - |
| *LDLR*: c.313+5G>A | 1 het |  | - |
| *APOB*:p.Ala444Val | 1 het |  | 0.00000958 |
| *APOB*:p.Val3857Met | 1 het |  | 0.0000581 |
| *APOB*:p.Val715Ile | 1 het |  | 0.00000205 |
| *APOB*:p.Arg2444Cys | 1 het |  | 0.0000212 |
| *APOB*:p.Val4171Met | 1 het |  | 0.00000616 |
| *APOB*:p.Glu3154Asp | 1 het |  | - |
| *APOB*:p.Phe3779Leu | 1 het |  | - |
| *APOB*:p.Lys518Asn | 1 het | 1 het] | 0.00000547 |
| *APOB*:p.Leu347Ile | 1 het |  | 6.84E-07 |
| *APOB*:p.Arg1689His | 1 het |  | 0.00174 |
| *APOB*:p.Leu3576Phe | 1 het |  | 0.0000041 |
| *APOB*:p.Pro1143Ser | 1 het |  | 0.00245 |
| *APOB*:p.Ser241Cys | 1 het |  | 0.00000616 |
| APOE:p.Glu88Gln | 1 het |  | 0.00000274 |
| *PCSK9*:p.Asp129Asn | 1 het |  | 0.00000821 |
| *PCSK9*:p.Val280Ala | 1 het |  | 0.00000137 |
| *PCSK9*:p.Asp37Asn | 1 het |  | - |
| *PCSK9*:p.L22_L23dup | 1 het | 2 het | 0.00327 |
| **BENIGN** |  |  |  |
| *LDLR*:p.Glu277Lys | 7 het | 2 het | 0.000457 |
| *APOB*:p.Leu21_Leu22dup | 20 het | 60 het and 2 hom | 0.000621 |
| *APOB*:p.Asp1113His | 5 het | 6 het | 0.00847 |
| *APOB*:p.Val4265Ala | 7 het | 2 het | 0.00893 |
| *APOB*:p.Arg1128His | 2 het | 2 het | 0.00463 |
| *APOB*:p.Val4128Met | 1 het | 6 het | 0.00722 |
| *APOB*:p.Ile4314Val | 1 hom | 4 het | 0.00788 |
| *APOB*:p.Pro994Leu | 1 het and 1 hom |  | 0.00106 |
| *APOB*:p.Ser3279Gly | 1 het | 4 het | 0.00325 |
| *PCSK9*:p.Leu23del | 1 het |  | 0.000228 |

**Table S3**

| **Gene** | **Variant** | **carriers FH cohort (n=300)** | **AF gnomAD** | **ClinVar** |
| --- | --- | --- | --- | --- |
| ***ABCG5*** | p.Met622Val | 3 het | 0.00606 | Benign/Likely benign |
|  | p.Tyr301His | 3 het | 0.00000274 | Uncertain significance |
|  | p.Ile244Thr | 1 het | 0.0000315 | Uncertain significance |
|  | p.Ala98Gly | 5 het | 0.00214 | Conflicting classifications of pathogenicity |
|  | p.Val524Ile | 2 het | 0.0000691 | Uncertain significance |
|  | p.Thr489Met | 1 het | 0.0000417 | not reported |
|  | p.Arg446Gln | 1 het | 0.0000164 | Conflicting classifications of pathogenicity |
|  | p.Asn440Lys | 2 het | 0.0000129 | Uncertain significance |
|  | p.Thr388Met | 1 het | 0.000163 | Conflicting classifications of pathogenicity |
|  | p.Arg105Leu | 1 het | - | not reported |
|  | p.Val47Phe | 1 het | 0.0003213 | benign |
|  | p.Gly27Ala | 1 het | 0.003959 | Conflicting classifications of pathogenicity |
|  | p.Gly10Arg | 1 het | 0.00001642 | Uncertain significance |
| ***ABCG8*** | p.Thr401Ser | 4 het | 0.0021 | Conflicting classifications of pathogenicity |
|  | p.Asp410Asn | 1 het | 0.0000129 | not reported |
|  | p.Glu85Asp | 1 het | 0.0000157 | Uncertain significance |
|  | p.Cys200Arg | 1 het | 0.00000342 | not reported |
|  | p.Arg263Trp | 1 het | 0.00000889 | Uncertain significance |
|  | p.Asp358Gly | 1 het | 0.00000342 | Uncertain significance |
|  | p.Asn409Ser | 3 het | 0.0000705 | Conflicting classifications of pathogenicity |
|  | p.Lys496Glu | 2 het | 0.0000411 | Uncertain significance |
| ***LPL*** | c.430-6C>T | 27 het and 1 hom | - | Benign/Likely benign |
|  | p.Ala427Thr | 2 het | 0.00138 | Benign/Likely benign |
| ***ABCA1*** | p.Gln2196His | 1 het | 0.000183 | not reported |
|  | p.Glu2093Asp | 1 het | 0.00000137 | not reported |
|  | p.Ala1182Thr | 1 het | 0.000882 | Conflicting classifications of pathogenicity |
|  | p.Val399Ala | 3 het | 0.00462 | Conflicting classifications of pathogenicity |
|  | p.Arg230Cys | 46 het and 1 hom | 0.00269 | Benign |
|  | p.Arg1680Gln | 1 het | 0.00054 | Conflicting classifications of pathogenicity |
|  | p.Val771Leu | 1 het | 0.000183 | not reported |
|  | p.Phe426Leu | 2 het | 0.0000424 | uncertain-significance |
|  | p.Arg174Gly | 2 het | 0.00000274 | not reported |
|  | p.Pro85Leu | 1 het | 0.00135 | Conflicting classifications of pathogenicity |
|  | p.Leu1408Phe | 1 het | 0.0000779 | Benign/Likely benign |
| ***LIPA*** | p.Lys254Met | 1 het | 0.00000205 | Uncertain significance |
|  | p.Arg127Gln | 1 het | 0.0000793 | Uncertain significance |
